# Status of Magnetic Resonance Imaging Systems and Quality Control Programs in Nigeria

**DOI:** 10.1101/2023.06.20.23290883

**Authors:** Maruf Adewole, Taofeeq A. Ige, Nicholas Irurhe, Philip Adewole, Michael Akpochafor, Ayo Ibitoye, Samuel Adeneye

## Abstract

Magnetic Resonance Imaging (MRI) employs the use of magnetic field and radio waves to produce images of the body. Quality Control (QC) is essential for ensuring optimal performance of MRI systems, as recommended by American College of Radiology (ACR), American Association of Physicists in Medicine (AAPM), and the International Society of Magnetic Resonance in Medicine (ISMRM). This survey examines the status of MRI systems and QC in Nigeria. Questionnaires were administered through google form to Radiologists, Radiographers, Medical Physicists, and biomedical engineers working in various MRI centers across the country, with a total of 44 responses received from 24 centers. The professional bodies of the professionals involved facilitated the questionnaire administration. The survey results indicate that 1.5T is the most common field strength of MRI systems in the country. 83% of the imaging centers rely solely on the service engineer to keep the MRI operational. Although 71% of the centers have Radiation Safety Advisors (RSA), their services do not include MRI. Moreover, 45% of the centers lack an understanding of the composition and importance of MRI QC. This is due to factors such as the absence of regulatory requirements, high patient workload, no trained personnel, and the unavailability of QC equipment. The findings of this survey highlight the need for improved QC programs in the country to improve image quality and longevity of MRI systems. It also underscores the need for the establishment of a regulatory framework and national policy to ensure the safe use of MRI in Nigeria.

## 1 Introduction

Magnetic Resonance Imaging (MRI) is a medical imaging technique that uses a powerful magnetic field, radio waves, and a computer to produce detailed images of the body’s internal structures. MRI is useful for diagnosing a variety of conditions, including brain and spinal cord injuries, tumors, joint injuries, and musculoskeletal disorders. It come in different strengths, measured in Tesla (T), with higher strength machines producing better image quality. MRI machines require routine maintenance and Quality Control (QC) to ensure they are functioning properly and producing accurate images. It encompasses a range of measures and procedures aimed at ensuring optimal performance. These measures include routine calibration, performance testing of MRI systems and strict adherence to protocols. The goal is to identify and prevent any issues that may compromise image quality and patient safety while ensuring compliance with the recommended standards and guidelines established by organizations such as American College of Radiology (ACR), American Association of Physicists in Medicine (AAPM), and the International Society of Magnetic Resonance in Medicine (ISMRM).

By conducting regular QC on MRI systems, healthcare facilities are able mitigate the risk of diagnostic errors, reduce the need for repeat scans, and ultimately enhance patient outcomes. Numerous standalone tools have been created to streamline and simplify routine QC testing, allowing for greater standardization, convenience, and time efficiency[1]–[5]. In addition, cutting-edge phantoms are currently under development to expedite QC procedures[6]

The emergence of MRI-guided radiation therapy has led to the integration of MRI units with LINACs, thereby increasing the need for rigorous QC of MR systems. In response to this demand, specialized tools have also been developed to effectively meet these requirements[7], [8].

The installation of the first MRI in Nigeria occurred in 1999 at the National Hospital Abuja. Since then, multiple units have been established throughout the country. According to a 2018 survey, there were 58 MRI units in the country[9], but this figure is now on track to reach nearly 100.

Medical Physicists bear the crucial responsibility of conducting QC on all imaging modalities within medical clinics. Unfortunately, this profession is currently experiencing a shortage of qualified personnel in the country[10].

By the guideline of ACR and AAPM, QC tests for MRI systems includes H0 static field homogeneity, Signal to Noise (SNR) and Contrast to Noise (CNR) measurements, Spatial Uniformity of SNR, Ghosting Ratio and MR image uniformity, Geometric Distortion and Spatial Linearity, Slice thickness, High Contrast Spatial Resolution and Low Contrast Object Detectability. These test has since been confirmed to be a veritable tool in ensuring optimal functionality of MRI systems [11], [12].

The paucity of research on survey of QC for MRI systems in Nigeria, and in contrast to similar investigations conducted in other countries [13]–[15] underscores the need for the present study. By exploring the state of MRI systems with respect to QC in Nigeria, we aim to shed light on an underexplored facet of medical imaging technology in the region. Through this inquiry, we hope to gain a comprehensive understanding of the current situation of MRI centers in the country and establish a baseline for future research and initiatives aimed at improving QC procedures in this context.

## 2 Materials & Methods

The survey was conducted through the utilization of a Google form, a web-based survey platform, for its ease of administration and data collection. To ensure a comprehensive and inclusive survey, the questionnaire was shared with the professional associations of Radiologists, Radiographers, Biomedical Engineers and Medical Physicists in Nigeria, whose members are key stakeholders in the field of medical imaging. The survey was open for a period of four months, from September to December 2022, to allow for adequate time for responses to be gathered. The duration of the survey was carefully chosen to balance the need for obtaining a significant number of responses with the need to minimize the burden on the respondents.

The questionnaire used in the survey consists of Thirty-two (32) questions structured into five distinct sections. The first section focused on collecting demographic information from the respondents, including details about their professional background, the hospitals where they worked, and the types of radiology equipment present in those hospitals.

The second section of the questionnaire evaluated the process of equipment procurement, with specific emphasis on the level of input from end-users during the procurement process. This section sought to determine if end-users were consulted in the procurement process and if their input was taken into consideration.

The third section of the questionnaire centered on the conduction of pre- and post-installation tests for equipment certification. The aim was to assess the level of equipment suitability for the end-users and the extent to which the equipment met the desired functions.

The fourth section of the questionnaire focused on the presence of quality Control measures and planned preventive measures within the hospital setting. This section aimed to determine if quality Control programs were in place and if preventive measures were being taken to avoid equipment breakdown and downtime.

The last section of the questionnaire explored the possible reasons for breakdown of equipment, the process of equipment repair, and the causes of downtime for broken-down equipment. This section aimed to gather insights into the factors that contributed to equipment failure and how hospitals dealt with such situations.

## 3 Result & Discussion

Forty-four (44) responses were received from 24 unique centers. These respondents consist of 52.3% (n =23) Radiologists, 43.2% (n = 19) Radiographers, 2.3% (n =1) Biomedical Engineer and 2.3% (n = 1) Medical Physicists. Multiple responses from same centers were aggregated to form 24 unique responses. The data from these responses can be inferred to represent the situation in other centers not captured in this survey.

**Figure 1:**
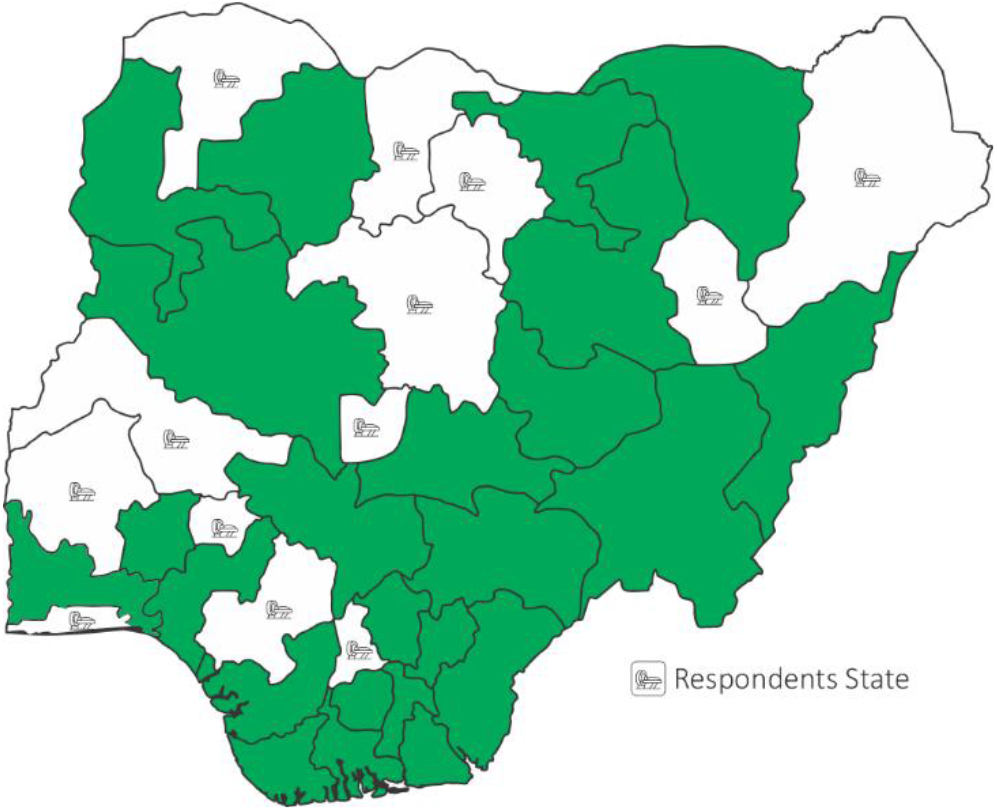
Distribution of Respondents location across Nigeria.

**Figure 2:**
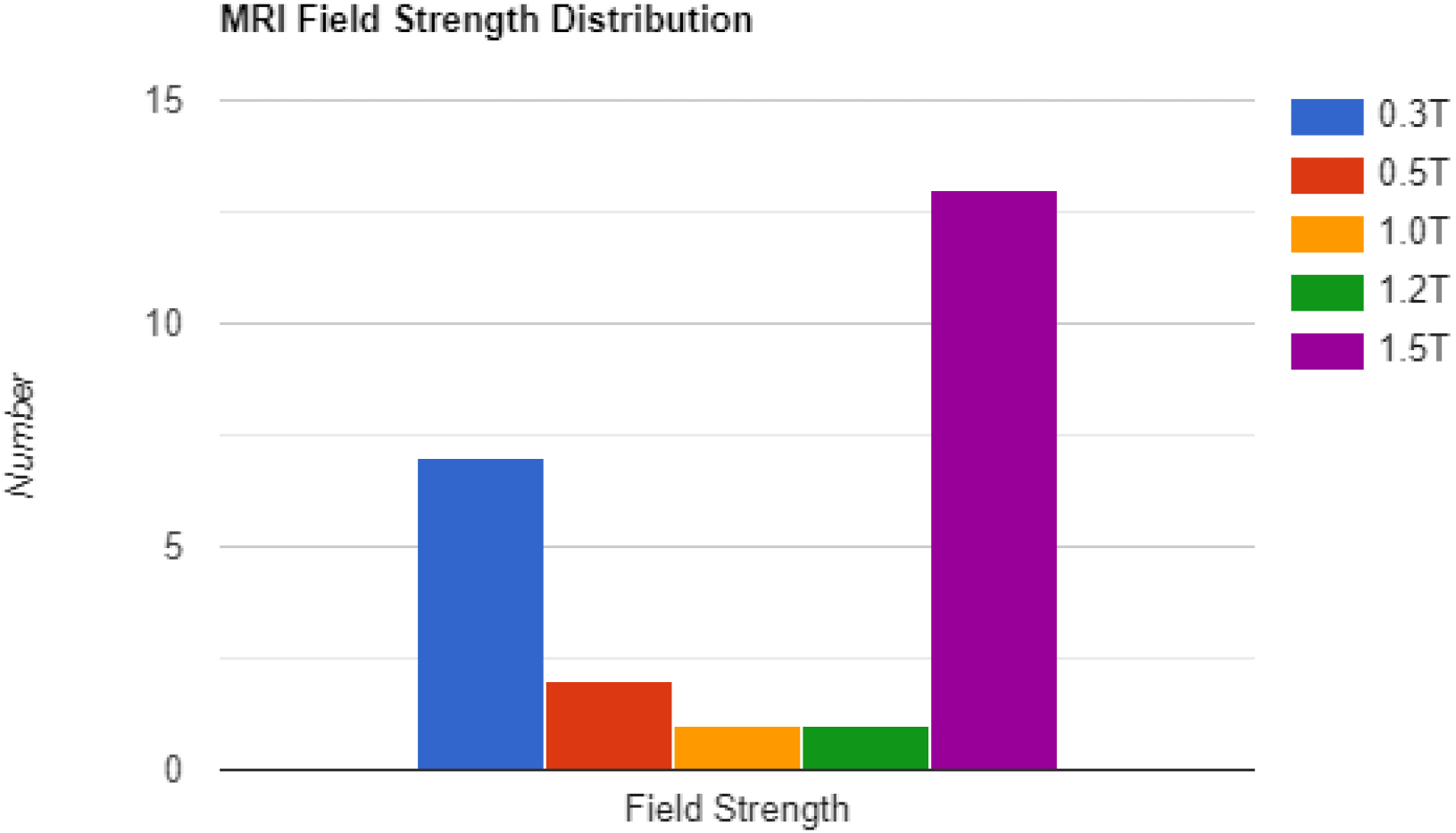
MRI Field Strength Distribution.

1.5T accounts for more than 50% of MRI machines from the responses. Most of the 1.5Ts were installed in the last 5 years. This shows that 1.5T is slowly becoming the field strength of choice in the country in place of 0.3Ts which most have been in service for more than 10 years.

Government owned centers tend to be busier than their colleagues with an average workload of 18 patient/week, Public Private Partnership (PPP) centers follows closely with average of 15 patient/week while solely Private owned centers have a workload of 8 patients/week.

**Figure 3:**
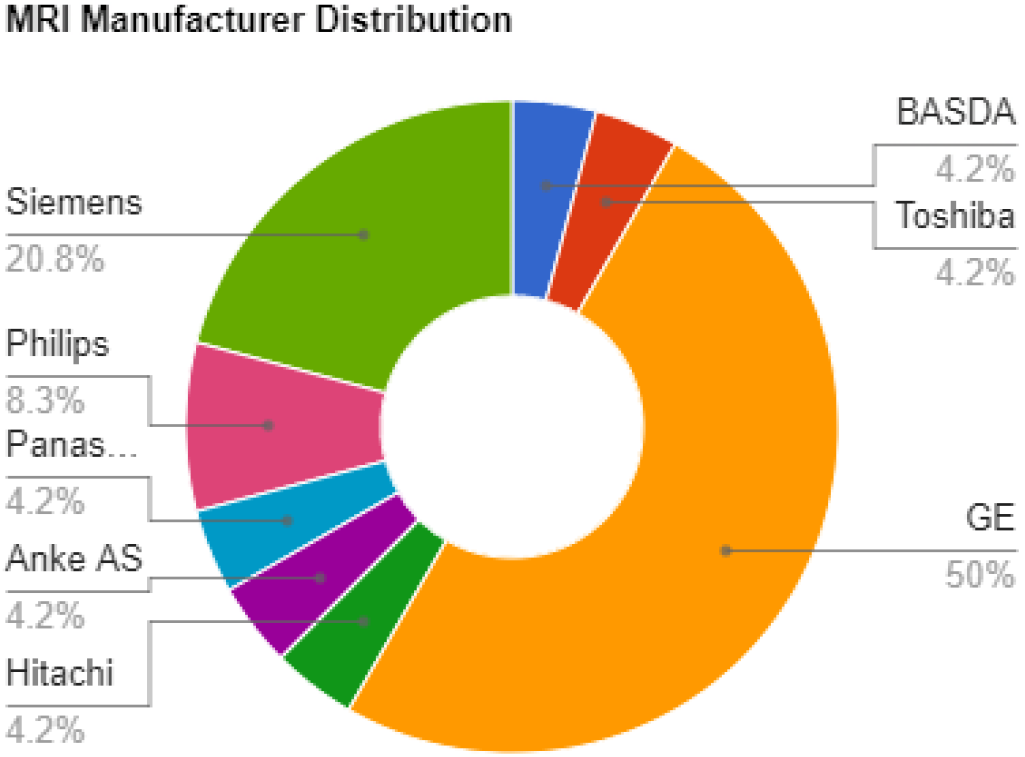
Distribution of MRI OEMs in Nigeria.

GE machines tend to dominate the OEM space with 50% installed in respondents centers closely followed by Siemens at 20%. Most of these centers (58%) have Pictures and Archiving Systems (PACS).

Majority of the center do not have an in-house Medical Physicist (63%). The few that have, are government hospitals with half of them for running radiotherapy services.

Almost all (71%) of the centers engage the services of Radiation Safety Advisor. This is because of their other imaging modalities that employ the use of ionizing radiation. This is a licensing condition mandated by the Nigerian Nuclear Regulatory Agency (NNRA). Conduct of routine QC on these other imaging modalities is in place almost all the centers (88%).

More than half of the centers confirm that acceptance testing was performed on their machine before the commencement of clinical operations. However, the acceptance is often performed by the Biomedical Engineer from the equipment vendor that supplied and installed the machine. This also include the RF cage assessment.

**Figure 4:**
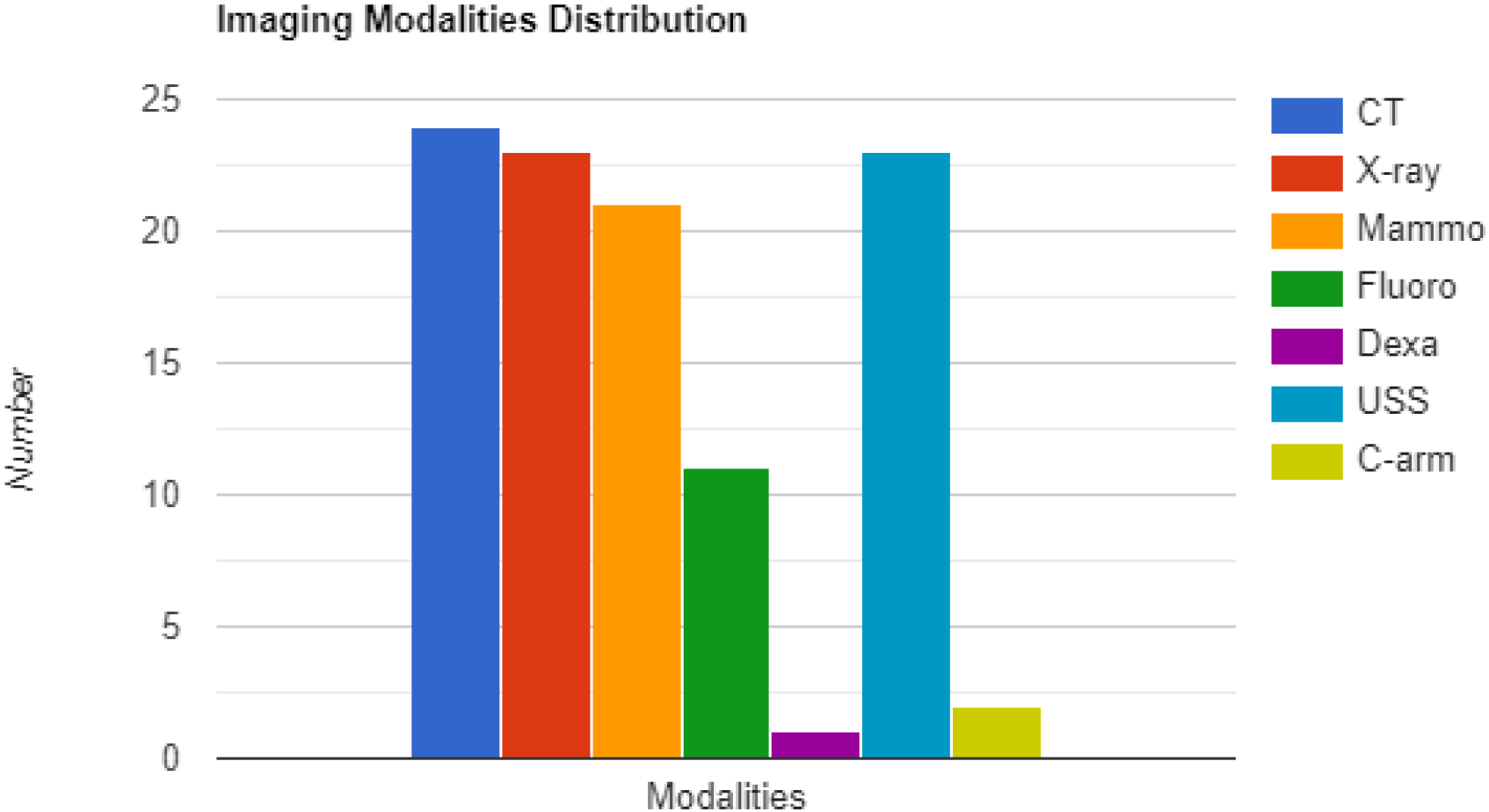
Available Imaging Modalities in Respondents Centers.

All centers that responded to this survey have CT (n = 24) in addition to their MRI. Xray and Ultrasound are the next modality of choice that was combined with MRI with (n = 23) likewise Mammography at (n = 21). Very few centers have Dexa (n = 1) and C-arm (n = 2). This is likely due to the fact that C-arm typically finds application in the theatre and not a routine radiology department. Fluoroscopy also has mild acceptance with (n = 11)

Routine QC is performed in only 58% of the centers, daily warm-ups are performed in only 62%. Of the few that perform routine QC, 42% performs it quarterly, 25% does it monthly while the remaining 33% doesn’t have a scheduled QC program.

Despite the foregoing, these centers take MRI safety very seriously, all of them placed appropriately signs such as “NO METALLIC OBJECT ALLOWED” at the entrance of the Faraday cage. A whopping 85% of respondents (n = 21) also believed that QC is important. 58% of respondents attests that downtime of MRI system is not frequent and even when they do, the parts that are most associated with causing it are Air conditioning and Chillers, Power outages and The RF Generator.

**Figure 5:**
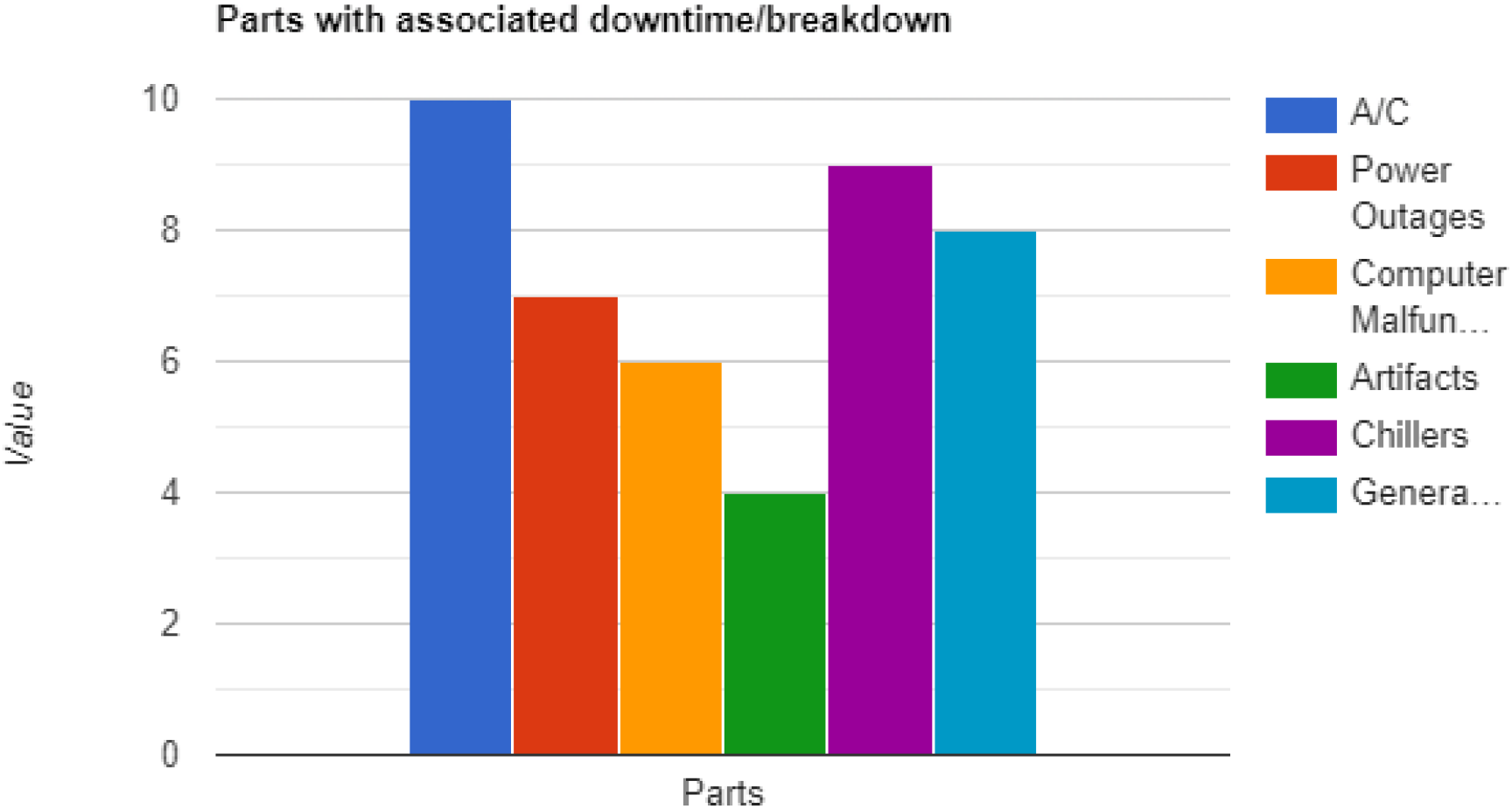
Distribution of Parts that are usually associated with downtime.

To forestall downtime, most centers (63%) have engaged the services of Biomedical engineers to perform routine preventive maintenance services at least once in a quarter. This is in agreement with the 83% of the respondents who believed that Preventive Maintenance is important. However, they do not share this believe at the same level with QC. This might explain why only half of the respondents believed QC is necessary after major repairs.

From table 1 above, it shows that some QC tests are performed more frequently than others. Screening patients for metallic objects, magnetic field homogeneity, and helium gas level monitoring are performed in most centers. However, tests such as low contrast detectability and transmitter gain/attenuation are performed in fewer centers.

**Table 1:** Frequency of tests performed in Imaging centers.

| QC Test | Number of centers that perform it |
| --- | --- |
| Screen patient for metallic objects | 13 |
| Scan reject analysis | 5 |
| Mechanical checks | 10 |
| RF shield assessment | 9 |
| Helium gas level monitoring | 12 |
| Magnetic Field homogeneity | 13 |
| Low contrast Detectability | 4 |
| Signal Noise Ratio | 10 |
| High contrast spatial Resolution | 7 |
| Image intensity uniformity | 7 |
| Slice thickness accuracy | 8 |
| Slice position accuracy | 9 |
| Transmitter gain/attenuation | 5 |
| Image artifact assessment | 7 |
| Geometric accuracy | 7 |
| Ghosting | 7 |

**Table 2:** Reasons attributed for non-performance of QC.

| Reasons | Number of centers |
| --- | --- |
| None | 4 |
| Machine not functional | 1 |
| Management not interested | 4 |
| No personnel to perform QC | 9 |
| Personnel available but no training | 6 |
| Scanner unavailable due huge patient load | 4 |
| No QC equipment | 6 |

Several centers face challenges when it comes to performing QC tests. The reasons for not performing QC tests vary across centers. In some centers, there are no apparent reasons for not performing QC tests, while in others, machine functionality issues or scanner unavailability due to a heavy patient load are factors preventing the performance of tests. Additionally, some centers face issues related to management, where management may not prioritize QC testing.

One common issue in many centers is a lack of personnel available to perform QC tests. Some centers have personnel, but they lack the necessary training to conduct the tests effectively. Other centers may not have access to QC equipment, which is essential for performing accurate tests.

Only half of the respondents have an MRI safety program in effect in their centers as against Imaging protocols that are available in almost all the centers (83%).

The proper maintenance and adherence to safety guidelines in MRI (Magnetic Resonance Imaging) is crucial to ensure accurate diagnosis and patient safety. One essential step in achieving this is having an in-house Medical Physicist who can oversee the development and adherence to MRI QC (Quality Control) and safety guidelines[16]. This would help to identify potential issues and take appropriate corrective actions.

In Ghana, a similar survey was conducted to determine the number of available MRI machines in the country [13]. This information provided an understanding on the accessibility and distribution of MRI services in the country. Similarly, a survey carried out in Saudi Arabia revealed that the lack of legal requirements and over-reliance on preventive maintenance by equipment vendors discouraged the conduct of QC on MRI[14]. This is similar to the situation in most centers in Nigeria too. However, such vendors are not responsible for conducting QC. This highlights the need for regulatory bodies to establish guidelines and standards to ensure the proper maintenance and QC of MRI machines in the country.

Another survey performed in the United Kingdom emphasizes the need to have a well-defined scope and frequency for MRI QC programs[17]. This concept would ensure that the necessary tests are performed regularly, and any potential issues are identified and addressed promptly.

In addition, it has been observed that in some clinics, end-users are not adequately involved in the procurement of radiodiagnosis machines[15]. This could lead to issues such as inadequate training and maintenance of the machines, which can impact the quality of diagnosis and patient safety.

In summary, this report discusses the results of a survey of MRI centers in Nigeria. The survey reveals that 1.5T MRI machines are becoming the field strength of choice, with GE machines dominating the OEM space. Most centers do not have an in-house medical physicist, but nearly all engage the services of an RSA. Routine quality Control (QC) is not performed in most centers, but centers take MRI safety seriously, with all of them placing signs such as “NO METALLIC OBJECT ALLOWED” at the entrance of the Faraday cage. Lack of personnel is a significant reason for not performing QC tests. Preventive maintenance is considered important, but QC is not seen as necessary after major repairs.

## 4 Conclusion

The number of MRI machines in the country keeps increasing due to increased population and clinical needs. These newly installed machines feature advance technology and increased field strength. These machines are owned by various entities such as the government, public-private partnerships, and private entities. However, there is a concerning lack of attention given to quality Control (QC) for MRIs. Several reasons contribute to this, including the fact that MRIs are not currently regulated by the NNRA, and management at MRI centers may not prioritize QC. Additionally, there may be insufficient personnel and equipment to conduct QC, and a lack of awareness regarding the importance of QC. It is crucial to address these issues and ensure that QC for MRIs is given the attention it deserves. To maintain the safety and accuracy of these diagnostic tool, it is essential that there is an increased interest in the QC of MRI. This includes the owners of MRI machines, as well as the personnel handling them. Improving the quality of image produced by MRIs and extending the life of the machines can be achieved through proper QC measures. Therefore, it is crucial that all stakeholders understand the importance of QC and take appropriate steps to ensure that it is implemented effectively.

## Data Availability

All data produced in the present study are available upon reasonable request to the authors

## Acknowledgement

The Authors would like to express their sincere gratitude and appreciation to all the respondents who generously dedicated their time and effort to complete the survey form. Their valuable participation has been instrumental in enriching the research and contributing to the findings presented in this manuscript.

## Notes

### Competing Interest Statement

The authors have declared no competing interest.

### Funding Statement

This study did not receive any funding

